# Integrating novel biomarkers and physiology in monitoring Fontan patients: FGF23, HAOX1 and arterial wave reflection

**DOI:** 10.64898/2026.05.11.26352952

**Authors:** Rada Ellegård, Asma Gul, Joanna Hlebowicz, Petru Liuba, Cecilia Gunnarsson, Constance G Weismann

## Abstract

Patients with Fontan circulation face evolving risk for cardiovascular morbidity and mortality, yet the interplay between cardiac function, vascular properties, and circulating proteins is incompletely defined. We hypothesized that biochemical biomarkers and multimodal cardiovascular profile differ significantly between Fontan patients and controls, and that selected markers may serve as predictors of reduced single ventricle function. We conducted a prospective observational study at a tertiary pediatric heart center including 31 individuals with Fontan circulation and 52 matched controls. Cardiac function was assessed by echocardiography; vascular phenotyping included carotid intima–media thickness, central and peripheral blood pressure, augmentation index corrected for heart rate, carotid–femoral pulse wave velocity, aging index, and reactive hyperemia index.

Compared to controls, the Fontan group had increased pulse wave reflection and central systolic pressure as well as decreased echocardiographic markers of systolic and diastolic function, while pulse wave velocity and other vascular parameters were not significantly different between the groups. Levels of 92 circulating cardiovascular biomarkers were quantified in a subset of 25 of the Fontan cohort and 81 controls using a proximity extension assay. Twenty-two biomarkers differed significantly in the Fontan group compared to controls, including FGF23, REN, HAOX1, and IL17D. Levels of several of these biomarkers correlated with patient age. Most importantly, HAOX1 (a peroxisomal oxidase linked to redox metabolism) and FGF23 (a bone-derived hormone regulating phosphate and vitamin D homeostasis) correlated negatively with ejection fraction within the Fontan group. By contrast, BNP was not associated with cardiac function in the Fontan group. None of the biomarkers correlated with central arterial parameters. In summary, central arterial hemodynamics and biomarkers such as FGF23 and HOAX1 may improve monitoring of cardiovascular function in single ventricle patients with Fontan circulation.

## 1. Introduction

Congenital heart defects (CHD) are the most common type of birth defects. In a severe subset of CHD that affects between 2 and 8 in 10,000 live births ^1^, children are born with only one pumping chamber, i.e. a functional single ventricle. This condition is often fatal without intervention, but a series of palliative surgical procedures allows survival into adulthood for many patients. In patients with single ventricle physiology following palliation with a total cavopulmonary connection (TCPC) resulting in a “Fontan circulation”, the pulmonary and systemic circuits are connected in series ^2^. Fontan patients are at risk for both heart failure with preserved and/or reduced ejection fraction (i.e. HEFpEF and/or HEFrEF), particularly as they become adults ^3,4^. As there is no subpulmonary ventricle, the flow in the pulmonary circulation is non-pulsatile and depends on a low pulmonary vascular resistance as well as good diastolic function of the receiving single subsystemic ventricle. If diastolic cardiac function is impaired, filling of the systemic ventricle is reduced, resulting in reduced cardiac output and exercise performance even though the ejection fraction may be preserved. In adult Fontan patients with reduced diastolic function, arterial stiffness is also increased ^5^. Our hypothesis was that vascular function is impaired and biomarkers are elevated in Fontan patients even at young age, and that there is a correlation with decreased cardiac function.

In this study, we used a multimodal approach to assess the Fontan circulation at a late postoperative time point, ranging from assessment of cardiac and vascular structure, stiffness, and endothelial function to circulating cardiovascular biomarker levels. Our aim was to gain insights into the underlying pathobiology of the Fontan circulation on a physiological and molecular level and ultimately develop novel strategies for clinical monitoring of these patients.

## 2. Material and methods

### 2.1 Study Design and Participants

Thirty-one Fontan patients and 52 matched healthy controls age 8-35 years were included in this prospective observational study, which was conducted at the Pediatric Heart Center at Skåne University Hospital in Lund. Exclusion criteria were severe valvar stenosis or insufficiency in the sub-systemic ventricle, heart transplant, uncontrolled hypertension, other known relevant systemic or genetic disorders. Patients with failing Fontan circulation were excluded. 57 patients who met eligibility criteria were contacted. 31 (54%) patients participated in the study. All patients underwent cardiac and vascular phenotyping, and cardiovascular markers were assayed using proteomics in 25 of these.

### 2.2 Cardiac and Vascular Phenotyping

The study protocol has been described previously and included documentation of demographic variables, intima media thickness (cIMT), central and peripheral blood pressure, augmentation index corrected to a heart rate of 75 beats per minute (AIx75), carotid-femoral pulse wave velocity, as well as aging index and reactive hyperemia index (RHI) as a marker of endothelial function ^6^.

Ventricular systolic function was assessed using echocadiography (Epiq 7, Philllips, Netherlands) by measuring fractional area change as well as 3D ejection fraction, global longitudinal and circumferential strain using the software package for the morphologic right or left ventricle as appropriate (TomTec Imaging Systems, Unterschleissheim, Germany). In addition, inflow pulse Doppler and tissue Doppler (TDI; average medial and lateral E’) were used for assessment of diastolic function. Common carotid artery distensibility, stiffness index and strain were determined as well in analogy to methodology described previously ^6^.

### 2.3 Proteomic Sampling and Assay

92 cardiovascular markers were assessed in 25 of the 31 patients described above, as well as 81 controls. Proteins were quantified with Olink’s Proximity Extension Assay (Target 96 Cardiovascular II, Olink Proteomics, Uppsala Sweden), hereafter referred to as CVD II.

### 2.4 Ethical Statement

The study was approved by Lund University’s Ethics Committee (DNR #2017-243) according to the declaration of Helsinki. All participants and/or their legal guardians provided written informed consent.

### 2.5 Statistical analysis for vascular parameters

Categoric data are presented as number (%) and continuous variables as median (interquartile range). Differences between groups were tested using the Mann-Whitney-U test for numeric data and Chi-square for categorical data. Associations between variables were analyzed using the Pearson’s correlation test. Linear regression analyses correcting for relevant covariates were performed. Differences between groups were interpreted as significant if p <0.05. Statistical analysis was performed using the Statistical Package for Social Sciences (IBM SPSS, version 28, Chicago, Illnois).

### 2.6 Statistical analysis for proteomics

Biomarker levels are reported as Normalized Protein eXpression (NPX, log^2^). Olink’s internal incubation, extension, and detection controls and inter-plate controls were used for standard plate normalization. All samples passed Olink QC (no failures; some warnings only which did not trigger sample exclusion). Proteins with low call-rate (>20% < LOD) were excluded; <LOD values were imputed as LOD/√2. Inter-plate effects were corrected by Olink bridge normalization.

Differences in levels of circulating cardiovascular biomarkers were assessed with ANOVA, with a linear model comparing the control group and Fontan group to each other adjusted for age and sex. Significance was defined as a Benjamini-Hochberg adjusted p-value <0.05. Correlation between biomarker levels and physiological cardiovascular parameters was performed using Spearman, with Benjamini-Hochberg for false discovery rate (FDR) adjustment. Adjustment for covariables was performed using partial correlation (Spearman). Adjusted p values are reported (p_adj_). Correlation between biomarker levels and sex, systemic ventricle involvement and ACEi/ARB use was performed using Wilcoxon rank-sum test.

## 3. Results

### 3.1 Cohort and Conventional Measures

Thirty-one Fontan patients and 52 matched controls with a median age of 20 (IQR 12-22) years were included in the assessment of cardiac and vascular structure, stiffness, and endothelial function. There was no significant difference between age and sex of the Fontan and control groups (Table 1). Fontan patients tended to have a shorter height and had a significantly lower weight compared to the controls. Accordingly, body surface area was significantly lower, and there was a trend towards a lower BMI in the patient group. Systolic and diastolic blood pressure were not significantly different between the groups. However, heart rate was significantly higher, and all parameters of systolic and diastolic systemic ventricular function were significantly more abnormal in the Fontan group.

**Table 1:** Descriptive statistics comparing Controls to Fontan patients. Continuous variables are presented as median (interquartile range). Groups were compared using the Mann-Whitney test. Significance level (p) is provided.

|  | Controls (n=52) | Fontan (n=31) | P |
| --- | --- | --- | --- |
| **Demographics** |  |  |  |
| Age [years] | 21 (12 – 25) | 15 (12 - 22) | 0.100 |
| Male | 58% (30) | 71% (22) | 0.251 |
| Height [cm] | 172 (155-181) | 163 (156-175) | .087 |
| Weight [kg] | 70 (42-80) | 54 (42-68) | .021 |
| BMI [kg/m<sup>2</sup>] | 22.6 (18.2-24.3) | 19.4 (17.2-22.7) | .062 |
| BSA [m<sup>2</sup>] | 1.8 (1.3-2.0) | 1.6 (1.3-1.8) | .022 |
| Systolic BP [mmHg] | 115 (108-121) | 114 (109-126) | .668 |
| Diastolic BP [mmHg] | 66 (63-73) | 69 (63-73) | .658 |
| Heart rate [beats per minute] | 61 (52-72) | 70 (59-79) | .010 |
| **Echocardiographic characteristics** |  |  |  |
| 3D ejection fraction [%] | 62 (60-65) | 48 (42-55) | <0.001 |
| Global longitudinal strain [%] | -31 (-33--30) | -18 (-24--15) | <0.001 |
| Global circumferential strain [%] | -31 (-30 --33) | -18 (-15 --24) | <0.001 |
| Fractional area change [%] | 40 (38-47) | 36 (31-42) | <0.001 |
| AV-valve E/A | 2 (1.6-2.7) | 1.3 (.9-1.8) | <0.001 |
| Average TDI E' [cm/s] | 14.8 (13.4-16.5) | 7.3 (5.7-9.8) | <0.001 |
| Average TDI E/E' | 5.7 (5-6.4) | 11 (7-14.4) | <0.001 |
| **Vascular characteristics** |  |  |  |
| Common carotid artery |  |  |  |
| - Distensibility | 0.007 (0.006-0.009) | 0.008 (0.005-0.0084) | 0.711 |
| - Stiffness Index | 3.4 (2.7-3.9) | 3.2 (2.9-4.2) | 0.939 |
| - Strain [%] | 11.5 (9.9-12.8) | 12.5 (8.6-14.2) | 0.458 |
| cIMT [mm] | 0.45 (0.42-0.49) | 0.44 (0.41 – 0.48) | .546 |
| Central Systolic Pressure [mmHg] | 99.8 (93.0-105.0) | 102.5 (95.9-109.6) | .126 |
| Central Diastolic Pressure [mmHg] | 67.0 (63.5-73.5) | 70.5 (64.5-75.1) | .522 |
| cAix75% | -11.8 (-17.6--3.1) | 9.8 (5.0-14.6) | <.001 |
| pAix75% | -22.0 (-28.0--13.5) | -5.0 (-11.0--2.0) | <.001 |
| Aging Index | -0.8 (-1.0--0.7) | -0.5 (-0.7--0.4) | <.001 |
| Vascular age [years] | 29.5 (23.0-36.0) | 42.0 (35.0-46.1) | <.001 |
| Pulse Wave Velocity [m/s] | 5.8 (4.3-6.8) | 5.3 (4.7-6.2) | .972 |
| Reactive Hyperemia Index | 1.8 (1.6-2.4) | 2.0 (1.5-2.6) | .640 |

**Table 2.** Characteristics of the Fontan Cohort, continuous variables expressed as median (IQR) and categorical variables expressed as number (%).

| Characteristics | Fontan Cohort (n=31) |
| --- | --- |
| Age at Fontan operation (years) | 2.1 (1.8-2.7) |
| Time since Fontan (years) | 13.5 (10 - 20.4) |
| Extracardiac Fontan, n (%) | 24 (77%) |
| Ventricular Morphology, n (%) |  |
| - Right | 18 (58%) |
| - Left | 13 (42%) |
| Medication n (%) |  |
| - Warfarin | 4 (13%) |
| - ASA | 26 (87%) |
| - ACE or A2R inhibitors* | 9 (29%) |
| History of Fontan associated complications# n (%) | 7 (23%) |
| Very good or good health (subjective assessment by patient) n (%) | 24 (77%) |

\*Angiotensin converting enzyme or Angiotensin 2 receptor inhibitor.
Data are expressed as number (percent) or median (interquartile range) as appropriate.

### 3.2 Vascular and Hemodynamic Phenotyping

In terms of vascular characteristics, all measures of common carotid artery elasticity were similar between the groups. Likewise, there was no statistically significant difference in *c*IMT or central systolic and diastolic blood pressure. Interestingly, though, both central and peripheral augmentation index corrected to a heart rate of 75 per minute as well as aging index were significantly higher in the Fontan group. By contrast, pulse wave velocity, a measure of general, large, arterial stiffness, as well as reactive hyperthermia index were not significantly different between the groups (**Table 1**).

### 3.3 Adjusted Analyses and Age Associations

The results remained unchanged following correction for relevant covariates (age, sex, height and heart rate), except that central systolic pressure was significantly higher in the Fontan group (B=4, 95% CI 0.1-8.4, p=0.046). Within the Fontan cohort, age correlated negatively with diastolic function (E’ -0.434, p 0.015), but not with parameters of systolic function. In addition, age correlated with central systolic pressure (r 0.364, p 0.048), and cIMT (r 0.463, p 0.009), but not with any of the other vascular parameters. None of the vascular parameters correlated with systolic or diastolic cardiac function within the Fontan cohort. A subgroup analysis comparing patients with single right vs. left ventricular morphology revealed no statistically significant differences between the two groups, though there was a trend towards lower fractional area change and ejection fraction in the systemic right ventricle group (p=0.068 and 0.095, respectively).

### 3.4 Circulating Cardiovascular Biomarkers

Circulating cardiovascular biomarkers were assayed in 25 of the Fontan patients and 81 controls of similar age. EDTA-plasma samples were collected at a late post operative timepoint, median 14 years (6-30 years).

PLS-DA ( Partial Least Squares Discriminant Analysis) analysis indicated differences in cardiovascular biomarker expression between the Fontan patients and the controls (**Figure 1A**). Of the 92 markers assayed, 22 were found to differ significantly between the control group and Fontan group when adjusted for age and sex (**Figure 1B-C, Suppl. Figure 1, Table 3)**.

**Figure 1.**
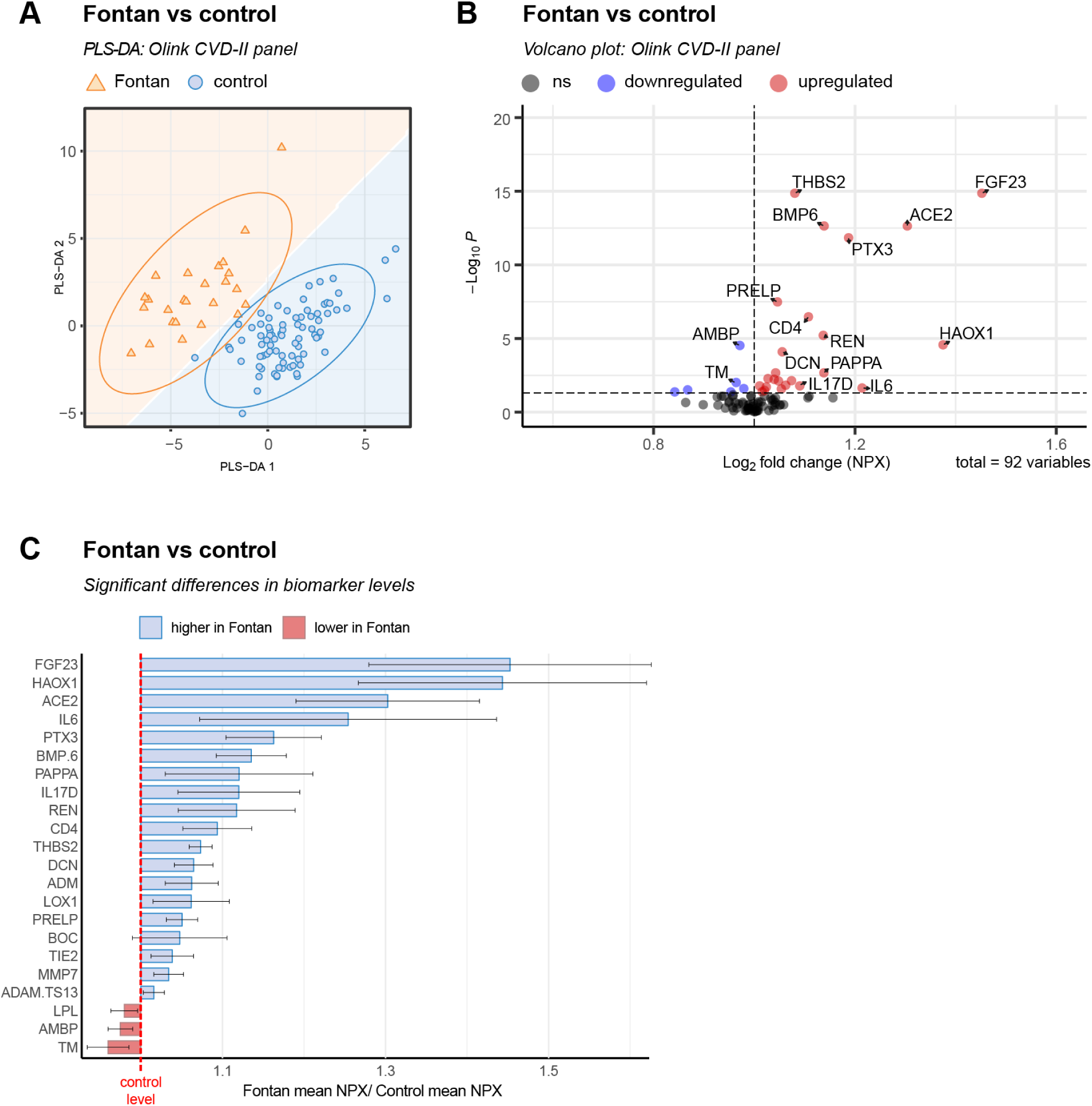
Circulating cardiovascular biomarkers were assayed in 25 Fontan patients and 81 controls using Olink CVD II. **(A)** Partial Least Squares Discriminant Analysis (PLS-DA) of expression levels in Fontan vs control was performed to examine if there are differences between the two groups. **(B)** Volcano plot of results from analysis of variance (ANOVA) with a linear model comparing the control group and Fontan group to each other adjusted for age and sex. Significance was defined as a Benjamini-Hechberg adjusted p-value <0.05. **(C)** Levels of biomarkers found to significantly differ in Fontan patients normalized to control. Bars how mean levels (normalized protein expression, NPX) in Fontan divided by mean levels (NPX) in controls. Error bars show 95% CI.

ACE2, REN, ADM, LOX-1, TIE2, ADAM-TS13, BMP-6, THBS2, PRELP, MMP7, DCN, PTX3, IL6, CD4, IL-17D, HAOX1, FGF23, PAPPA, and BOC levels were significantly higher, whereas TM, LPL, and AMBP levels were significantly lower in the Fontan cohort. The biomarkers displaying the most pronounced significant difference between Fontan and controls were HAOX1 (estimate=1.72, p_adj_ =<0.0001), and FGF23 (estimate=1.32, p_adj_ =<0.0001) **(Figure 1C)**.

One patient, who reported fever within 6 weeks of sampling, displayed markedly elevated levels of IL6, one patient displayed elevated ADAM-TS13 and two patients had elevated REN (**Suppl. Figure 1, Suppl. Figure 2**).

The correlation of biochemical markers with age and sex differed between the Fontan and control groups **(Figure 2).** BOC (ρ=-0.43, p_adj_ =0.020) and PTX3 (ρ=-0.46, p_adj_ =0.016) displayed significant negative correlation with age in the control group, and BOC (ρ=-0.55, p_adj_ =0.035), FGF23 (ρ=-0.61, p_adj_ =0.013) and CD4 (ρ=-0.62, p_adj_ =0.013) displayed significant negative correlation with age in the Fontan group. While PTX3 and CD4 correlated in the same direction in both groups (despite not reaching significance in both), FGF23 levels showed a different pattern. In the Fontan group, FGF23 had a significant negative correlation with age, while there was no discernible correlation in the control group. No markers were significantly associated with sex after FDR adjustment, in either group.

**Figure 2.**
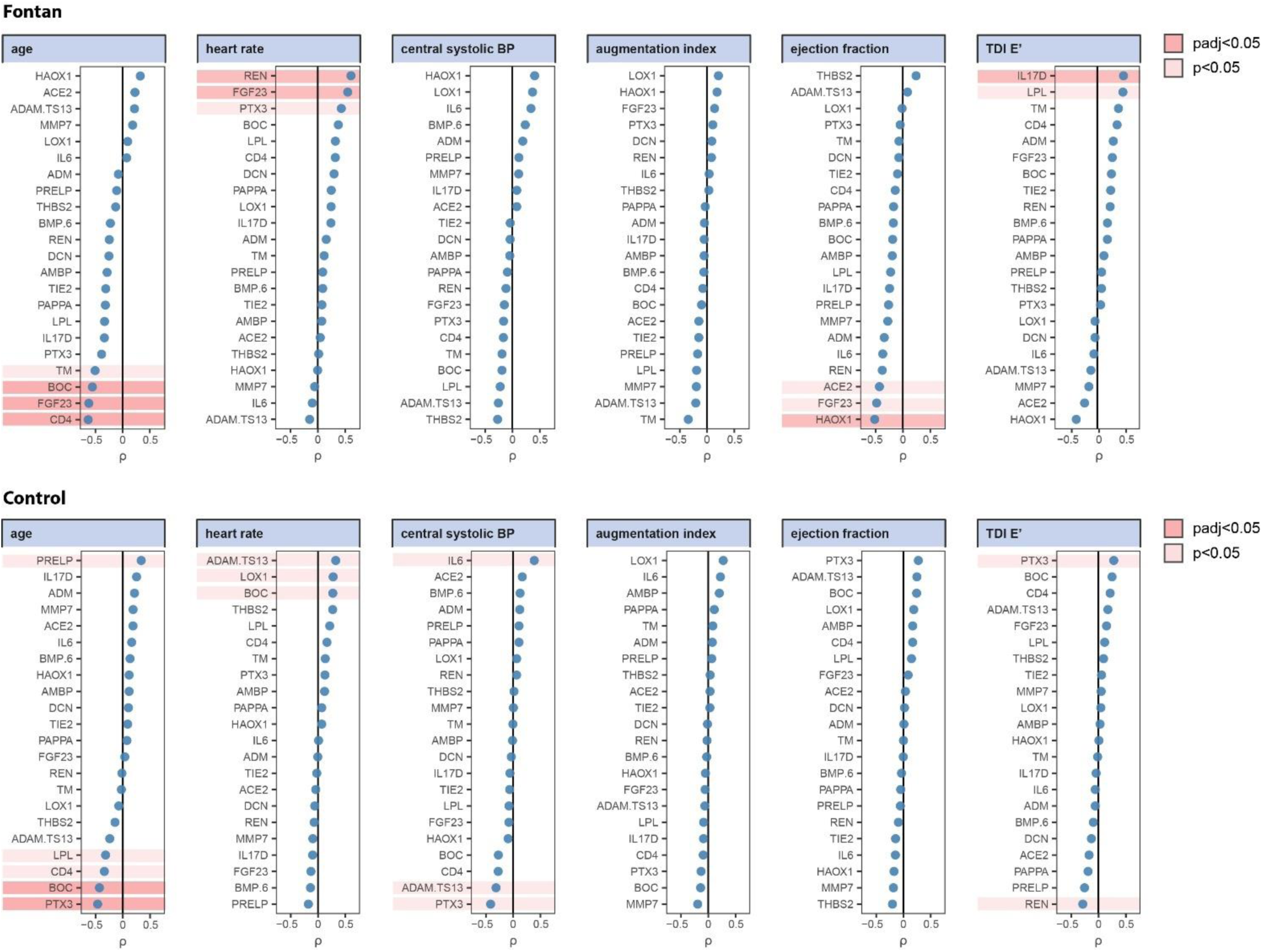
Correlation between biomarkers and physiological cardiovascular parameters in the Fontan group. The levels of the biomarkers in the Fontan group (N=25) found to differ significantly from the control group (N=81) were correlated to patient age (years) and the physiological cardiovascular parameters that displayed significant differences between the groups, ie average TDI E’ [cm/s], ejection fraction (%), augmentation index corrected to a heart rate of 75 beats per minute (AIx75; %), central systolic blood pressure (mmHg) and heart rate (1/min). Correlation was performed for the Fontan group only, using Spearman followed by Benjamini-Hechberg for FDR adjustment.

In the Fontan group, 11 patients had a systemic left, and 14 had a systemic right ventricle. There was no significant correlation between systemic ventricle type (right vs left) and biomarker expression. 6 patients were on angiotensin-converting enzyme inhibitors (ACEi) or angiotensin receptor blockers (ARB). There was a significant correlation between ACEi/ARB use and higher levels of REN (ρ = 0.57, p_adj_ =0.025), but no other biomarkers showed significant correlations. In the control group, none of the biochemical markers evaluated correlated significantly with physiological cardiovascular parameters **(Figure 2).**

### 3.5 Correlation to physiological cardiovascular markers in the Fontan group

Next, we focused on biomarkers that were significantly different between the groups and performed correlation analyses with relevant parameters of cardiovascular function within the Fontan cohort **(Figure 2)**. Physiological cardiovascular parameters found to differ significantly between the Fontan and control groups were selected for correlation analyses with biomarkers, i.e. age, heart rate, ejection fraction, average TDI E’ (cm/s), central systolic blood pressure (mmHg), and augmentation index corrected to a heart rate of 75 beats per minute (AIx75).

Interestingly, ejection fraction had a moderate negative correlation with HAOX1 (ρ= -0.51, p_adj_ =0.038) and almost met statistical significance for correlation with FGF23 (ρ= -0.47, p_adj_ =0.054). Better diastolic relaxation, reflected by the average annular TDI e′ velocity, correlated positively with IL-17D (ρ=0.51, p_adj_ =0.038). Higher heart rate significantly correlated with higher REN (ρ=0.60, p_adj_ =0.042) and FGF23 (ρ=0.54, p_adj_ =0.049).

### 3.6 Comparison with Published Fontan Proteomics

Literature studies identified two previous publications that had assayed similar biomarkers in Fontan patients as in our study– a study by Frank et al assaying infants with single ventricle heart disease undergoing Stage 2 palliation ^7^, and a cohort of adults with Fontan circulation studied by Kelly et al ^8^. A comparison between the results of these studies and ours is displayed in **Table 3**.

**Table 3.** Differences in circulating biomarkers between Fontan and healthy controls in this study compared to two previous publications. In this study, differences in circulating biomarkers were assayed in 25 Fontan patients and 81 similar age controls at a late post-operative time point using Olink CVD II panel. Two previous publications have assayed similar biomarkers in Fontan patients – Frank et al assayed biomarkers using Olink CVD II in 29 infants with single ventricle heart disease undergoing Stage 2 palliation and 25 similar age healthy controls, and Kelly et al used Olink Explore 3072 (which includes all markers in CVD II) to assay 58 adults with Fontan circulation and 29 controls of similar age. The biomarkers found to be significant in this study were compared to results from the two previous publications. As Frank et al. did not report a direct post-op vs control contrast; we derived it on the NPX (normalised protein expression, log2) scale as log2FC(post-op vs control) = log2FC(post-op vs pre-op) + log2FC(pre-op vs control) from their supplementary fold-changes, without recalculating p-values.

| This study (Fontan vs Control) |  |  | Frank et al. 2021 | Kelly et al. 2025 |
| --- | --- | --- | --- | --- |
| Protein | Pathway / Cluster | Direction (↑/↓) | Direction (↑/↓) | Direction (↑/↓) |
| ACE2 | Heart / RAAS<br>(neurohormonal strain) | ↑ | ns | ↑ |
| REN |  | ↑ | ns | ↑ |
| ADM |  | ↑ | ↑ | ns |
| LOX1 | Endothelium /<br>Coagulation | ↑ | ns | ns |
| TM |  | ↓ | ns | ns |
| TIE2 |  | ↑ | ns | ns |
| ADAM-TS13 |  | ↑ | ns | ns |
| BMP-6 | ECM remodeling /<br>Fibrosis | ↑ | ns | ns |
| THBS2 |  | ↑ | ns | ↑ |
| PRELP |  | ↑ | ↓ | ns |
| MMP7 |  | ↑ | ns | ns |
| DCN |  | ↑ | ns | ns |
| PTX3 | Inflammation<br>(innate/adaptive) | ↑ | ns | ns |
| IL6 |  | ↑ | ↑ | ↑ |
| CD4 |  | ↑ | ns | ns |
| IL17D |  | ↑ | ns | ns |
| HAOX1 | Lipid handling /<br>Oxidative stress | ↑ | ↑ | ns |
| LPL |  | ↓ | ↑ | ns |
| FGF23 |  | ↑ | ↑ | ↑ |
| PAPPA | Mineral & Endocrine<br>regulation | ↑ | ↑ | ns |
| AMBP |  | ↓ | ↓ | ns |
| BOC | Development &<br>Mechanotransduction | ↑ | ns | ns |

Of the 22 markers that differed significantly between the Fontan and control groups in the study presented herein, two – FGF23 and IL6 – showed corresponding significance in both other studies. Five markers (ACE2, REN, ADM, THBS2, and HAOX1) showed corresponding significance in one or both of the publications by Frank et al and Kelly et al ^7,8^, while 12 were non-significant in both comparator studies. Three markers showed conflicting results compared to least one prior study: PRELP was increased in our Fontan cohort, but decreased in Frank *et al* and non-significant in Kelly et al; LPL was decreased in our cohort, but increased in Frank et al and non-significant in Kelly et al; and TM was decreased in our cohort, but non-significant in both Frank et al and Kelly et al.

### 3.7 Data Availability

The data in this article can be shared upon reasonable request to the corresponding author.

## 4. Discussion

To our knowledge, this is the first study of Fontan patients that has evaluated biomarkers in the context of arterial and cardiac function using multiple modalities. We found evidence of altered central hemodynamics and impaired systolic and diastolic cardiac function, but no correlation between those factors. Conversely, we did identify HAOX1 and FGF23 as potential markers of systolic function in Fontan patients. In the following sections, we will first discuss physiologic parameters and then turn our attention to biomarkers and potential future clinical use thereof.

In terms of arterial characteristics, we found increased central systolic pressure in the Fontan group following correction for relevant covariates, reflecting an increased workload on the heart and other central organs such as the kidney ^9^. Fontan patients also had increased arterial wave reflection (augmentation and aging index) compared to healthy controls. Due to arterio-ventricular interaction, increased central arterial stiffness can have a negative impact on diastolic and systolic cardiac function ^6,10^. Single ventricle patients are at particular risk for both systolic and diastolic cardiac dysfunction ^11^. The lack of a correlation between arterial characteristics and cardiac function in the Fontan cohort can likely be explained by the heterogeneity of the different types of single ventricle type of CHD itself, whether the great arteries are normally related or transposed, aortic arch geometry, aortic wall stiffness, and prior surgeries such as aortic arch reconstruction or aortic coarctation repair ^12,13^. As pulse wave reflection and central systolic pressure increase with age and are known to adversely affect diastolic function in general, we suggest monitoring and treating central hemodynamics in addition to brachial blood pressure in Fontan patients.

The Fontan circulation is associated with increased venous pressure, resulting in lymphatic and liver congestion, and a pro-thrombotic tendency ^14–16^. In terms of biochemical markers, routine monitoring of Fontan patients includes NT-proBNP (brain natriuretic peptide) as well as routine laboratory tests such as liver function tests. Other markers such as MELD-XI, vWF/ADAMTS13 balance and sCD40L, have been used on a research basis and have not entered the clinical arena ^14,15,17,18^. Recent studies have suggested that additional markers may provide early signs of circulatory failure and be used to identify patients at risk for adverse outcomes ^19^. To identify additional biomarkers with clinical potential for monitoring Fontan patients, we measured 92 cardiovascular proteins using the Olink CVD II panel, which included biomarkers for endothelial health, coagulation/thrombosis, inflammation/remodelling, and cardiac stretch^20,21^. Of those, 22 proteins were differentially expressed between the Fontan and control groups, with HAOX1 and FGF23 displaying the most pronounced significant differences. Our study describes a distinct proteomic and neurohormonal signature that reflects the systemic physiological cost of the Fontan circulation.

FGF23, a hormone that controls phosphate and vitamin-D balance, emerged as one of two candidates for future clinical use. FGF23 is upregulated during renal stress and systemic venous congestion ^22^ and has been shown to be involved in pathological changes triggered by pressure or volume overload in the heart - although the exact molecular mechanisms behind this remain to be elucidated ^23^. A growing number of clinical studies show that elevated circulating levels of FGF-23 are associated with left ventricular hypertrophy (LVH), heart failure, and mortality - especially in patients with impaired renal function ^24^. Clinically, elevated FGF23 has been associated with heart-failure risk, systemic venous congestion, right ventricular dysfunction independent of BNP, left ventricular systolic dysfunction, cardiac fibrosis, atrial arrhythmias and renal dysfunction - all highly relevant to patients with Fontan circulation ^25,26^. As such, using FGF23 as a prognostic biomarker in clinical practice has been proposed ^25^ ^27^. The possibility to use FGF23 as a therapeutic target remains to be determined.

In our late post-operative Fontan cohort, FGF23 was elevated, which is in concordance with two prior studies on single ventricle patients - pre as well as post Fontan palliation ^28, 7,8^. FGF-23 upregulation is typically only observed in individuals >60 years of age, where it is associated with cardiovascular disease ^29^, and there were no differences in FGF23 levels attributable to age in our control group. Of note, in addition to being significantly upregulated in the Fontan group, FGF23 also had a significant positive correlation with heart rate, a negative correlation with age and nearly met adjusted significance level for correlation with ejection fraction within the Fontan group. None of these correlations were observed in the control group.

The seemingly paradoxical negative correlation of FGF23 and age may be explained by TCPC-specific changes in FGF23 expression over time. Following TCPC surgery, an acute change in hepatic venous pressure occurs. Over time, Fontan-associated liver disease (FALD) emerges, i.e. hepatic fibrosis and cirrhosis with age ^30^. In analogy, a mouse model evaluating FGF23 expression has shown that acute hepatic injury causes a significantly higher fold-increase in hepatic FGF23 expression compared to chronic fibrosis ^31^. Thus, young Fontan patients may be subjected to more active hepatic remodelling that later in life, where fibrosis is manifest.

The negative correlation of FGF23 with ejection fraction did not quite meet statistical significance (p=0.054), likely due to p-adjustment for multiple comparisons (22 biomarkers were differentially expressed on a group level). In the context of the published literature on FGF23, and keeping in mind the relatively small number of Fontan patients in our cohort, we still consider FGF23 a strong candidate that deserves further investigation as a prognostic marker in Fontan patients. In addition, the significant positive correlation between FGF23 and heart rate in our cohort may be indicative of early-stage heart failure.

In our study, Renin (REN) was associated with higher heart rate and ACE inhibitor and /or ARB use. REN is released from the kidney and triggers the renin–angiotensin–aldosterone system (RAAS), a neurohormonal pathway regulating vascular tone and sodium–water balance; its correlation with heart rate underscores the profound preload dependency of the Fontan circulation ^14,32^. In the absence of a sub-pulmonary pump, these patients rely on heart-rate reserve and neurohormonal activation (including RAAS) to support systemic perfusion when stroke volume/preload is constrained ^14,33^. The correlation of REN and heart rate in our study is consistent with these findings.

The herein observed link between REN and FGF23 further suggests a maladaptive neurohormonal–bone feedback loop: RAAS hormones such as Angiotensin II and aldosterone increase FGF23 expression in bone (the physiological source of FGF23) ^34,35^. Conversely, FGF23 can suppress renal ACE2 expression, plausibly shifting the balance toward Ang II signaling ^36^. This ‘Renin–FGF23 axis’ may contribute to adverse myocardial remodeling, consistent with experimental evidence that FGF23 can directly induce cardiomyocyte hypertrophy via FGFR signaling ^37,38^.

The second major candidate biomarker associated with single-ventricular systolic dysfunction in our study was HAOX1. HAOX1, also known as glycolate oxidase, is a liver-enriched peroxisomal enzyme that catalyzes the oxidation of glycolate to glyoxylate with concomitant generation of H₂O₂, linking hepatic redox balance to peroxisomal metabolism ^39,40^. Importantly, even though HAXO1 transcripton is downregulated during hepatic stress, there is evidence that the release of this liver-enriched HAOX1 protein into plasma is increased during hepatocellular stress or injury. HAOX1 expression tracks with liver-related phenotypes, including intrahepatic fat change, steatotic liver disease risk, and improvement in liver enzymes following bariatric surgery ^41–43^. In general, a predominance of translational responses over transcriptional responses in early hepatic hypoxic stress has been reported ^44^.

In our study, HAOX1 exhibited an inverse correlation with ejection fraction. Importantly, our data concerns circulating HAOX1 protein rather than hepatic transcript abundance. In the Fontan circulation, chronically elevated central venous pressure and reduced cardiac output promote hepatic congestion, impaired portal inflow, and relative hypoxia of centrilobular hepatocytes ^45^. Thus, increased circulating HAOX1 protein may reflect hemodynamically mediated hepatic stress within the cardiohepatic axis ^17,45,46^. Thus, association between higher circulating HAOX1 and lower ejection fraction is consistent with worsening hemodynamics being accompanied by greater hepatic stress and circulating HAOX1 protein. The precise mechanism underlying its elevation in plasma in Fontan patients remains to be clarified.

Lastly, we identified IL17D (a non-canonical cytokine) as a potential biomarker for diastolic impairment. Interleukin-17D (IL-17D) is a comparatively understudied member of the IL-17 cytokine family. Rather than signaling through canonical IL-17 receptor complexes (e.g., IL-17RA-containing receptors used by IL-17A/F), IL-17D has been shown to bind CD93, establishing an IL-17D–CD93 signaling axis ^47,48^. Of note, diastolic function decreases with age in general. In our Fontan cohort, following age correction, there no longer was a significant correlation of IL17D and diastolic function. Thus, we speculate that IL17D is not an independent marker of diastolic function.

To contextualize our findings, we reviewed proteomic studies most similar to our approach and focused on two for head-to-head comparison of significant proteins. Frank et al. profiled serum from inter-stage infants with single-ventricle heart disease 24 h post-op using the same CVD II panel as in our study ^7^. By contrast, Kelly et al. analyzed plasma from adults with established Fontan circulation using Olink Explore 3072, which includes our CVD II targets ^8^. Of the 22 markers found to be significantly different between the Fontan and control groups in the current study, 7 showed corresponding significance in one or both comparator studies (including FGF23, HAOX1, REN and IL17D), 12 were not significant in any, and 3 showed conflicting findings. Differences in biofluid/compartment, age and circulation stage, and timing (acute peri-operative vs chronic baseline) likely explain why some markers diverge across studies. Nevertheless, this study supports the presence of biomarkers characteristic of Fontan circulation, which may hold future clinical potential. Several discordant markers are biologically plausible given age/stage, sample type, timing, and therapy effects.

This was a single-centre study of Fontan patients with heterogeneous single-ventricle types (systemic right and left ventricles). The complexity of CHD and altered loading, where normal values for two ventricles do not necessarily apply to single ventricles, limits the utility of echocardiographic indices of systolic and diastolic function, and the relatively small cohort increases the risk of type II error. The proteomics analysis was cross-sectional and used a targeted Olink Cardiovascular II panel reporting relative NPX, so results reflect associations at a single time point and require external validation and prospective linkage to outcomes. Standardised pre-analytical handling and Olink QC/normalisation were applied, yet residual confounding by clinical covariates (e.g., medications, time since Fontan, renal/ hepatic status) cannot be fully excluded.

In conclusion, Fontan patients demonstrate augmented arterial wave reflection with both diastolic and systolic impairment compared to controls. Plasma levels of FGF23 and HAOX1 emerged as markers with prognostic value for potential future clinical use. Together, the addition of those markers may improve risk stratification and improve medical management of Fontan patients.

## Data Availability

Data available upon reasonable request.

---

**Supplementary Figure 1.**
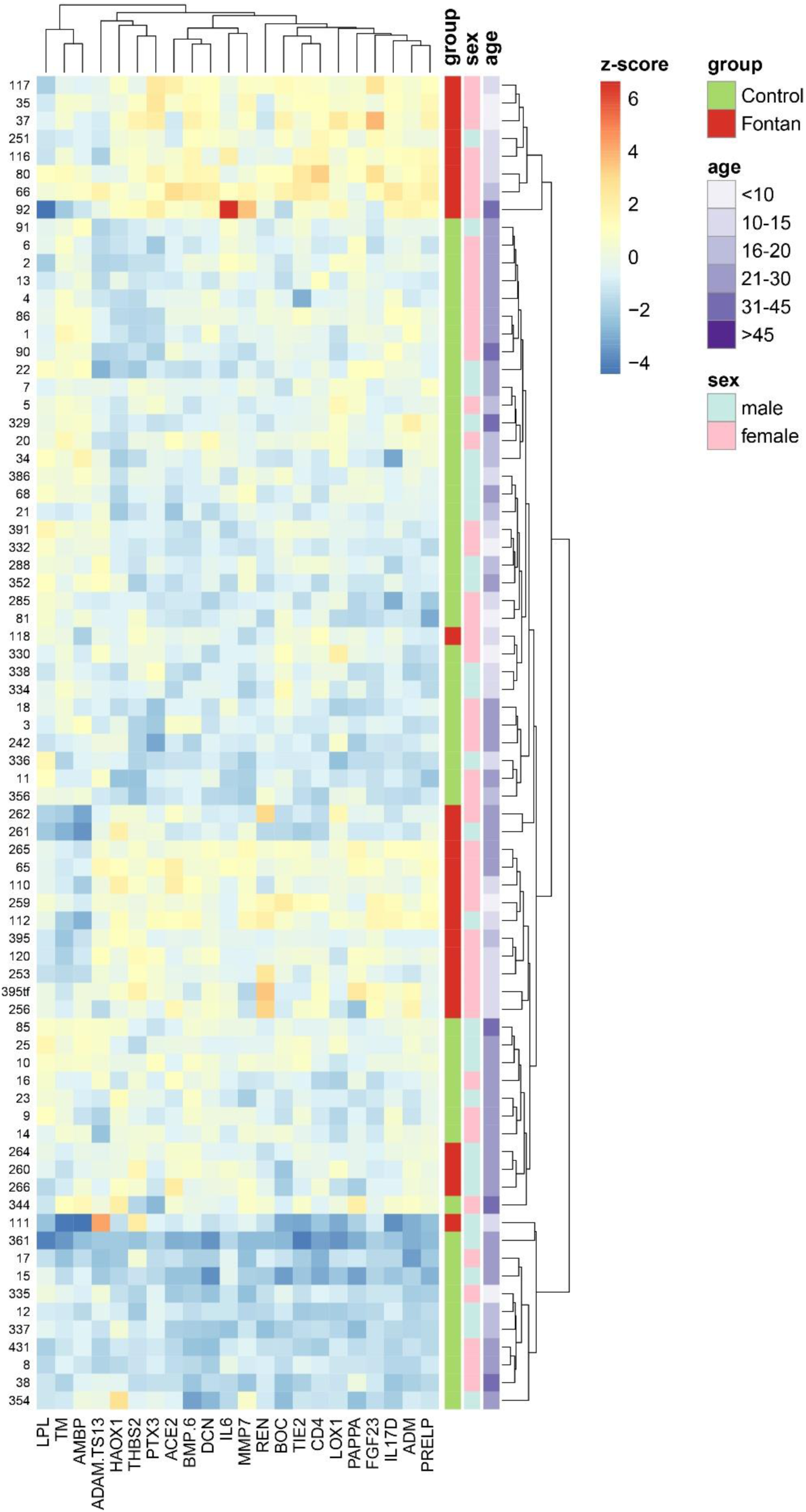
(Euclidean) Hierarchical clustering of levels of cardiovascular biomarkers found to differ significantly between Fontan and controls (after controlling for patient age and sex). Expression levels are shown as z-transformed NPX values.

**Supplementary Figure 2.**
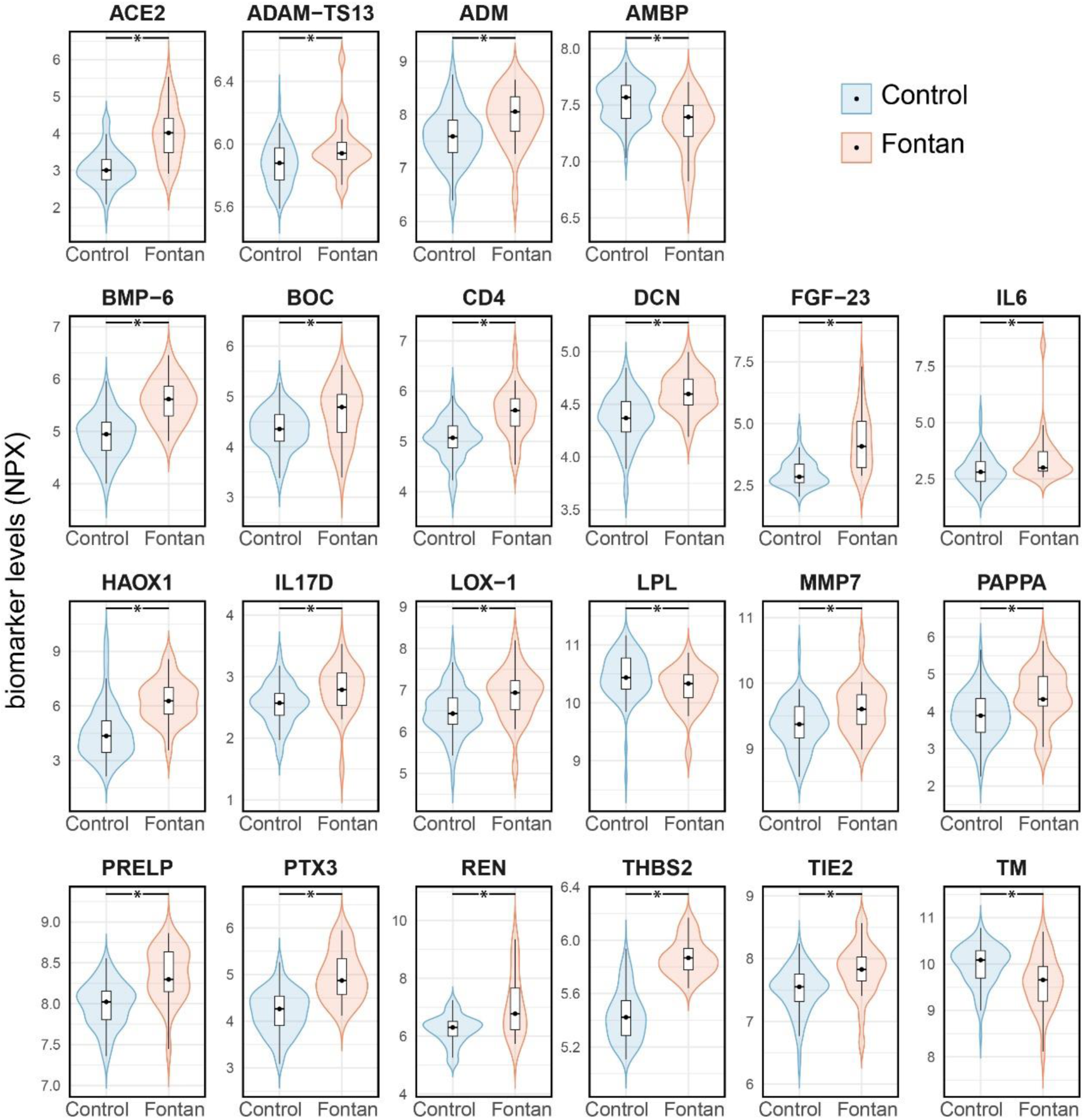
Levels (NPX) of cardiovascular biomarkers in Olink Cardiovascular II panel found to differ significantly between Fontan (N=25) and control (N=81) groups using ANOVA with a linear model adjusted for age and sex. Significance was defined as a Benjamini-Hechberg adjusted p-value <0.05.

